# Comparative Reconstruction of SARS-CoV-2 transmission in three African countries using a mathematical model integrating immunity data

**DOI:** 10.1101/2023.07.07.23292215

**Authors:** Bechir Naffeti, Walid BenAribi, Amira Kebir, Maryam Diara, Matthieu Schoenhals, Inès Vigan-Womas, Koussay Dellagi, Slimane BenMiled

## Abstract

**Objectives:** Africa has experienced fewer coronavirus disease 2019 (COVID-19) cases and deaths than other regions, with a contrasting epidemiological situation between countries, raising questions regarding the determinants of disease spread in Africa.

**Method:** We built a susceptible–exposed–infected–recovered model including COVID-19 mortality data where recovery class is structured by specific immunization and modeled by a partial differential equation considering the opposed effects of immunity decline and immunization. This model was applied to Tunisia, Senegal, and Madagascar.

**Finding:** Senegal and Tunisia experienced two epidemic phases. Initially, infections emerged in naive individuals and were limited by social distancing. Variants of concern (VOCs) were also introduced. The second phase was characterized by successive epidemic waves driven by new VOCs that escaped host immunity. Meanwhile, Madagascar demonstrated a different profile, characterized by longer intervals between epidemic waves, increasing the pool of susceptible individuals who had lost their protective immunity. The impact of vaccination in Tunisia and Senegal on model parameters was evaluated.

**Interpretation:** Loss of immunity and vaccination-induced immunity have played crucial role in controlling the African pandemic. Severe acute respiratory syndrome coronavirus 2 has become endemic now and will continue to circulate in African populations. However, previous infections provide significant protection against severe diseases, thus providing a basis for future vaccination strategies.

## 1. Introduction

Coronavirus disease 2019 (COVID-19) pandemic caused by the severe acute respiratory syndrome coronavirus 2 (SARS-CoV-2) coronavirus has been ongoing for over 3 years. According to World Health Organization (WHO) data^1^, the number of cases and deaths due to COVID-19 is significantly lower in Africa than in the rest of the world. However, lower testing rates, case detection rates, and deaths reported in Africa could account for the lower number of cases and deaths.

More importantly, SARS-CoV-2 has heterogeneously affected African countries (Naffeti et al., 2022). Tunisia, Senegal, and Madagascar reported 1, 152, 483, 86, 594, and 66, 098 confirmed cases and 29, 378, 1968, and 1403 deaths, respectively, by April 23, 2023. These disparities can be partly attributed to demographic factors. Compared to Senegal and Madagascar, Tunisia has a relatively older population demographic (median age, 32.8 years; life expectancy at birth, 76 years; individuals aged >65 years represent 8.86% of the population). These indicators were respectively 19 years, 62.5 years, and 3.1% in Senegal and 19 years, 57 years, and 3.47% in Madagascar, respectively^2^. The contrasting impact of the epidemic on African countries might also reflect differences in the severity and promptness of public health and social measures (PHSMs) imposed by the national health authorities to limit virus transmission. These PHSMs also shape the introduction and diffusion of new variants of concern (VOCs) into populations. In the past, African population might also have been diversely exposed to pathogens that share cross-reactivity to SARS-CoV-2, which conferred partial resistance to the emerging coronavirus. Finally, the genetic background and rate at which the immunity to SARS-CoV-2 wanes after natural infection may influence individual responses to the virus and resistance to reinfection. To the best of our knowledge, no previous study has reported the reconstruction of SARS-CoV-2 transmission in African countries using a mathematical model that integrates immunity data.

The humoral immune responses acquired after SARS-CoV-2 infection are typified by the production of antibodies against the viral nucleoprotein (N) and surface spike (S) protein (among other viral antigens). While antibodies to N protein are not protective (*i*.*e*. non-neutralizing), antibodies to S protein (and particularly to the receptor binding domain [RBD] domain) efficiently neutralize the excreted SARS-CoV-2 virus *i*.*e*. the original ancestral strain as well as the first VOCs (*i*.*e*. *α, β, δ* VOCs), but they are less effective against Omicron (Iyer et al., 2020; Romano et al., 2020; Gallagher and Buchmeier, 2001). Antibody titers to SARS-CoV-2 decline over time after infection and may fall below the threshold, usually within a few months [27]. The cellular immune responses mounted after SARS-CoV-2 infection also contribute to stronger resistance against severe disease after reinfection and likely account for an increasing cross-immunity over time observed in individuals who have experienced multiple reinfections (Yamayoshi et al., 2021). They may also account for the resistance to the virus in individuals who have lost detectable anti-S/RBD antibodies. All these features also apply to vaccine-induced immunity, with the vaccine type and its expressed protein(s) being additional factors.

In this study, we aimed to assess the role of immunity and viral variants in shaping the dynamics of the SARS-CoV-2 pandemic in Africa. Specifically, we conducted a comparative analysis with a focus on Tunisia, Senegal, and Madagascar, representing the Northern, Western, and Austral Africa, respectively. Our study incorporated epidemiological parameters including daily cases, daily deaths, epidemic waves, emerging VOCs, and the impact of the natural decay of post-infectious SARS-CoV-2 immunity within the population.

By investigating these factors, our study contributes to a comprehensive understanding of the SARS-CoV-2 pandemic in African countries, shedding light on the influence of immunity and viral variants on disease dynamics. The findings of this study can provide data for improving public health strategies and interventions tailored to the unique context of African countries.

## 2. Model description and parameters evaluation

We compared the SARS-CoV-2 pandemic dynamics in Senegal, Madagascar, and Tunisia using the susceptible–exposed–infected–recovered/Death (SEIR)/DS model developed by White and Medley (1998) and reviewed by Barbarossa et al. (2018) (Figure 1). The susceptible compartment (*S*) was divided into two groups: *S*_1_ for individuals who had previously been infected, identified by antibodies (Abs) to S/RBD but not to N, and *S*_2_ for those without prior infection (no N and no S/RBD Abs). Individuals in S_1_ or S_2_ could have been infected by exposed (*E*) or infected individuals (*I*), with E moving to I after developing antibodies (symptomatic or asymptomatic, respectively). I compartment was divided into *I*_*nd*_ and *I*_*d*_, where the former was not detected, and the latter tested positive. Individuals in the *I* group either recovered (*R*) or died (*D*).

**Figure 1:**
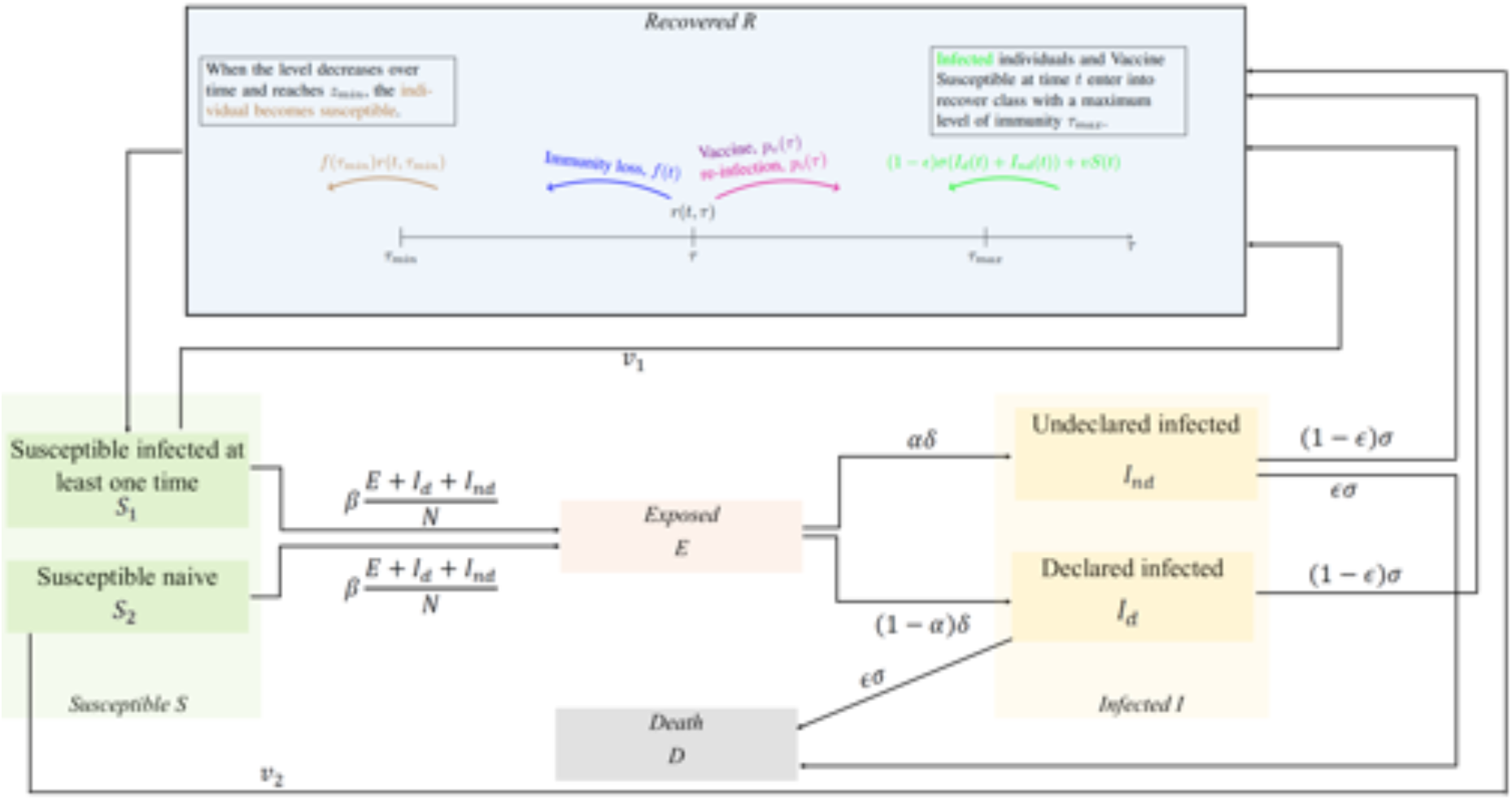
Conceptual diagram of the model.

We compared the SARS-CoV-2 pandemic dynamics in Senegal, Madagascar, and Tunisia using an SEIR/DS model developed by White and Medley (1998) and reviewed by Barbarossa et al. (2018) (Figure 1). The susceptible compartment (S) was divided into two groups: *S*_1_ for individuals who previously had been infected, identified by antibodies (Abs) to S/RBD but not N, and *S*_2_ for those without prior infection (no N ant no S/RBD Abs). Individuals in *S*_1_ or *S*_2_ can be infected by exposed individuals (*E*) or infected individuals (*I*), with *E* moving to *I* after developing Abs (symptomatic or asymptomatic people). *I* compartment is divided into *I*_nd_ and *I*_d_, where the former was not detected, and the latter were tested positive. Individuals in I either recover (*R*) or die (*D*).

We assumed that individuals who recovered from SARS-CoV-2, denoted as *R*, have different levels of residual immunity. We structured the compartments of recovered patients based on the levels of anti-S antibodies detected by enzyme-linked immunoassay, which is a good proxy for the levels of neutralizing antibodies. When an individual’s neutralizing antibody titer drops below a certain threshold, they are moved from the *R* pool to the susceptible (*S*_1_) pool. We used τ to denote the level of immunity (*i*.*e*. concentration of antibodies to S/RBD). Immunity loss was modeled using a partial differential equation, which accounted for both spontaneous decline and/or boosting through vaccines and/or exposure to the virus. The density of recovered individuals at time *t*, denoted as *r*(*t*, τ), was determined based on antibody level τ. *R*_*input*_(*t*) represented the influx of recovered individuals at each time unit, while *S*_*input*_(*t*) represented the flux of individuals who have lost their immunity. The total immunized population was obtained by integrating *r*(*t*, τ) over the range of τ values. More details are provided in appendices A and B and Table 1.

The increase in the number of infections led to an increase in the recovery class inflows, R. Each observed epidemic wave generated a cohort of individuals who became infected simultaneously and subsequently entered the recovery class as a group, creating a corresponding cohort wave in the seroimmune space, represented by *r*(*t*, τ). The seroimmune wave declined over time at a rate *f(t*) and ultimately lost after 1/*f*(*t*). To evaluate the kinetics of antibodies to the S or N proteins of SARS-CoV-2 [*i*.*e*. *f*(*t*)], we used four datasets acquired in Senegal between August 14, 2020 and August 17, 2021 (corresponding to epidemic waves 2, 3, and 4) (Talla et al., 2022). Data were normalized using technology and proteins. We divided the normalized kinetic data into four time groups: > 150 days; [50, 150*days*]; [23, 50*days*], and < 23*days*, and performed a linear regression analysis, called the linear kinetic model, and a logistic regression analysis, called the non-linear kinetic model. A similar analysis was performed for immunoglobulin G antibodies against the N protein (Appendix Table 2 and Figure 7).

The threshold below which recovered patients become susceptible was evaluated using a pre-pandemic database built in 2018 with sera from individuals who had never been infected with SARS-CoV-2 (Senegalese Dielmo 2018 cohort). We observed that the time required for immunity loss, *T*, falls within the range of 180λ 210 days, which is consistent with the findings of previous studies by Pollán et al. (2020); Dan et al. (2021). As the linear regression yielded an *R*2 > 0.8 for all *T* À [180, 210] days, we conducted a sensitivity analysis on *r*(*t*, τ) with respect to τ the range of 180 to 240 days.

We incorporated daily case and death data from WHO databases for Senegal, Tunisia, and Madagascar in our model^3^. The model parameters are listed in appendix Table 1. Vaccination data from Tunisia and Senegal were integrated into the model, while excluding data from Madagascar due to low vaccination coverage. External data sources, including the Oxford SARS-CoV-2 Government Response Stringency indexfootnote https://www.bsg.ox.ac.uk/research/covid-19-government-response-tracker, Google Mobility Index^4^^5^, and public health and social measures, PHSMs, were used to contextualize the epidemiological data curves and national public health decisions.

## 3. Results

Our model was simulated using COVID-19 pandemic data collected from Senegal (for both linear and non-linear kinetic models) and from Tunisia and Madagascar (for linear kinetic models) between March 2020 and September 2022. We used antibody levels against S/RBD as a proxy for SARS-CoV-2 immunity acquired after natural infection or vaccination.

The model exhibited a close match to the data on daily cases and daily deaths (Figures 2a, 2b, and 2c). Additionally, the simulated density of immune individuals, *r*(*t*, τ), in the pool of recovered individuals closely approximated the data from the first national seroprevalence surveys in Madagascar and Senegal (Figures 3g and 3h). The epidemic analysis in the three countries allowed us to distinguish between the cases of Senegal and Tunisia cases and the case of Madagascar.

**Figure 2:**
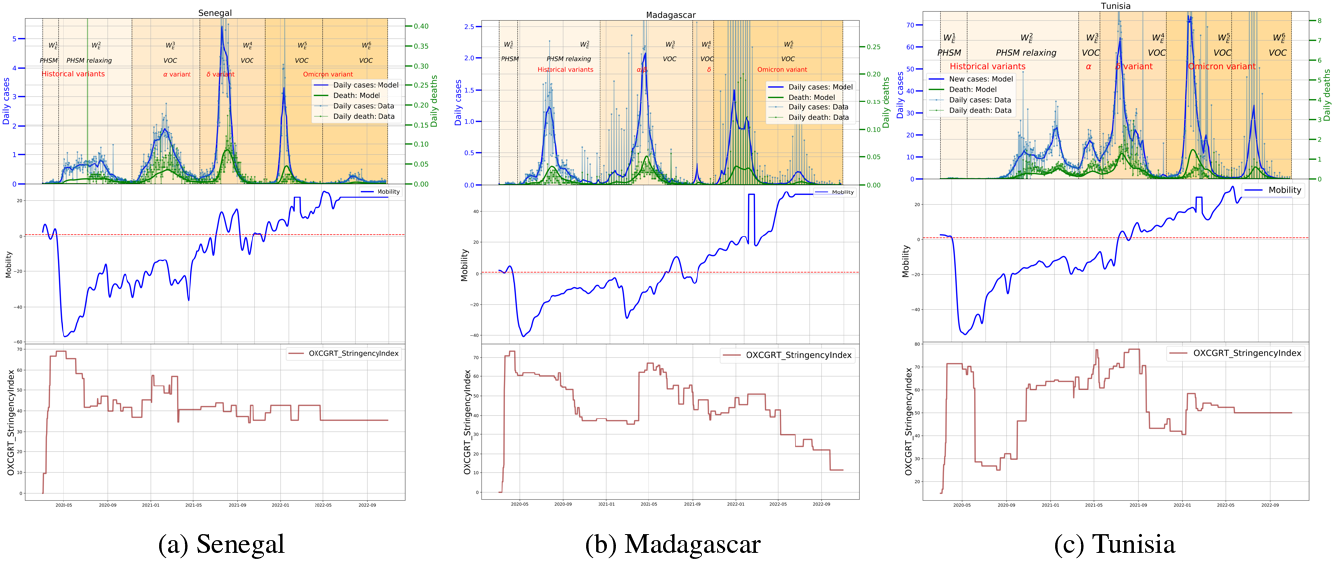
From left to right: Senegal 2a; Madagascar 2b; and Tunisia 2c. Each country is successively represented from top to bottom: the simulated data for daily cases, daily deaths, and daily hospitalizations per 100, 000 individuals curves, the Google mobility curve, and the Oxford stringency index curve from the beginning of the epidemic to Oct. 2022 The simulated data for infected individuals (*I*) fit both the daily cases of SARS-CoV-2 and reported data from national seroprevalence surveys. We observed that Senegal and Tunisia have experienced six waves, whereas Madagascar has experienced five waves. The three countries have undergone historical, *α, β*, and omicron VOCs. The mobility curves of the three countries were similar. Except for the first wave corresponding to the start of the epidemic, when severe preventive measures and PHSMs actions were effectively implemented, the two indicators are rising and will become positive between April and June 2021, indicating the relaxation of these actions in all countries. Except for the first wave, Senegal and Madagascar had very few cases according to the stringency index. Although significant PHSMs decisions have been made in Tunisia, they have not been implemented.

**Figure 3:**
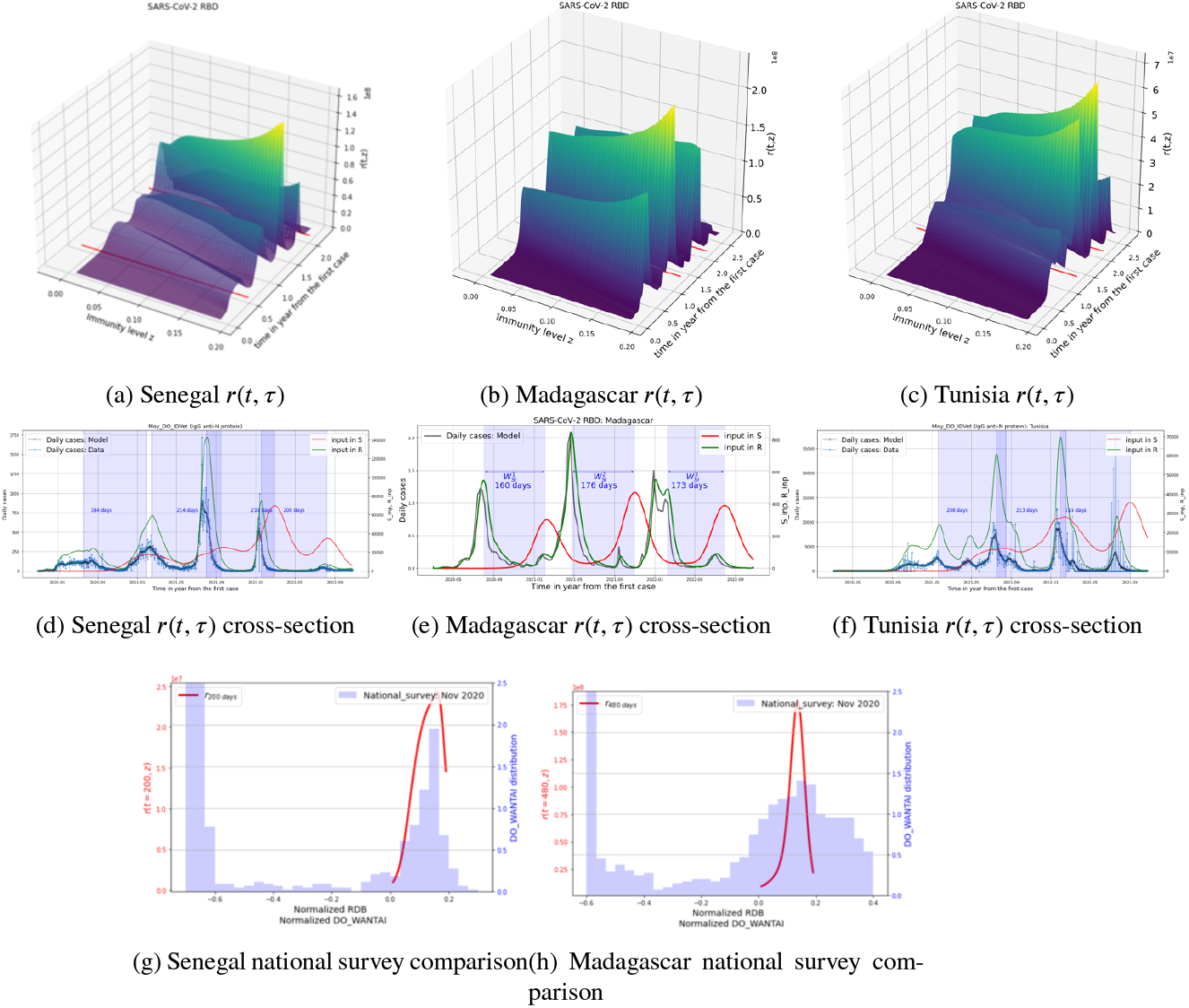
Figures 3a, 3b, 3c represent the simulation data of the density of recovered individuals at time *t* with seroimmune level τ, *r*(*t*, τfrom the start of the epidemic to September 2022 (Ì 2.5 years), with τ levels of concentration of antibodies to S/RBD. The values of *r*(*t*, τ) are represented per 100, 000 individuals. Figures 3d, 3e and 3f represent the cross-section of *r*(*t*, τ) for τ = τ_*max*_ (*R*_*inpu*t_ class) and τ = τ_*min*_ (*S*_*input*_ class) as well as the epidemic curve based on daily cases by time (in days) with daily cases according to field data (vertical blue bars). Figures3g and 3h compare r(t, τ) at the time of the national seroprevalence survey with data from the survey on antibodies against S/RBD (note that Tunisia did not measure antibodies against RBD during its national seroprevalence survey). The simulated data for the density of immune individuals (*r*(*t*, τ)) fit both the daily cases of SARS-CoV-2 and the reported data from national seroprevalence surveys. Seroimmune waves are generated by epidemiological waves. The subsequent epidemiological wave ended in Tunisia and Senegal. In Senegal, 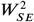 and 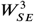 intersected slightly (Figures (Figures 3d and 3f)). In Madagascar, the seroimmune wave ended before the epidemiological wave (figure 3e). In Madagascar, a succession of independent seroimmune waves of a duration of 160 to 180 days was observed.

In Senegal and Tunisia, based on the mobility index, VOC distribution, and stringency index (Figure 2), the epidemic was divided into three successive phases. The first epidemic phase, *Ep*_1_, corresponded to epidemic wave 1 (*W E*_1_) caused by the historical virus entering the two countries through infected visitors. As the diffusion of the virus was constrained locally by strong public health measures and limited to early case contacts, *W E*_1_ was inconspicuous. The second phase, *Ep*_2_, corresponded to the epidemic 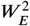 wave in Senegal and Tunisia (Figures 2a and 2c) caused by diverse lineages of the ancestral virus, but it was much more severe than *Ep*_1_ (BenMiled et al., 2023). The resumption of the epidemic in *Ep*_2_ could be ascribed to a relaxation of PHSMs, borders reopening during the tourism season, and the geographic extension of the epidemic to previously unaffected regions. The third phase, Ep_3_, corresponded to 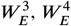,and 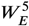 waves, characterized by the successive emergence ofα, δand Omicron VOCs in Tunisia and Senegal citepChouikha2022,Padane2022. These VOCs had a selective advantage over the preceding lineages and almost completely displaced them. This period also coincided with an almost complete drop in PHSMs, as indicated by a mobility index close to zero or positive (Figures 2a and 2c). As expected *δ* and Omicron waves were characterized by a faster virus spread and more deaths (Figure 2). We observed that the seroimmune waves *r*(*t*, τ) ended with a subsequent epidemic wave (Figures 3d and 3f). For instance, the second and third seroimmune waves (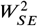 and 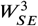 and 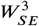 and 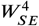, respectively) intersected to a certain degree (figures 3d and 3f). In Tunisia, an inconspicuous initial epidemic wave led to a barely perceptible initial seroimmune wave. The seroimmune waves resulting from the second and third epidemic waves completely overlapped and merged into a unique seroimmune wave, 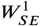, 208 days in duration (Figure 3f).

Finally, two phases are identified in the recovered (*R*) curve (Figures 4a and 4c). The first phase lasted from the start of the epidemic to months 9/10 of 2021, during which *R* increased in a stepwise manner, up to 45% and 55.65% respectively. The increase in R resulted from the overlap of successive seroimmune waves and accumulation of immune individuals. During this first phase, the pool of susceptible individuals, *S*, was mainly composed of individuals naive to SARS-CoV-2 (*S*_2_), while individuals who had previously been infected but lost their immunity and become susceptible again (S_1_) were relatively fewer. Thus, *S*_1_ and *S*_2_ were 13.59% and 40.84%, respectively, in Tunisia and 16.86% and 25.86%, respectively, in Senegal (Figure 4a). The second phase in the *R* curve encompassed epidemic waves 5 and 6 caused by the Omicron, characterized by a steep decrease in *R* due to a significant immunity loss in the population.

**Figure 4:**
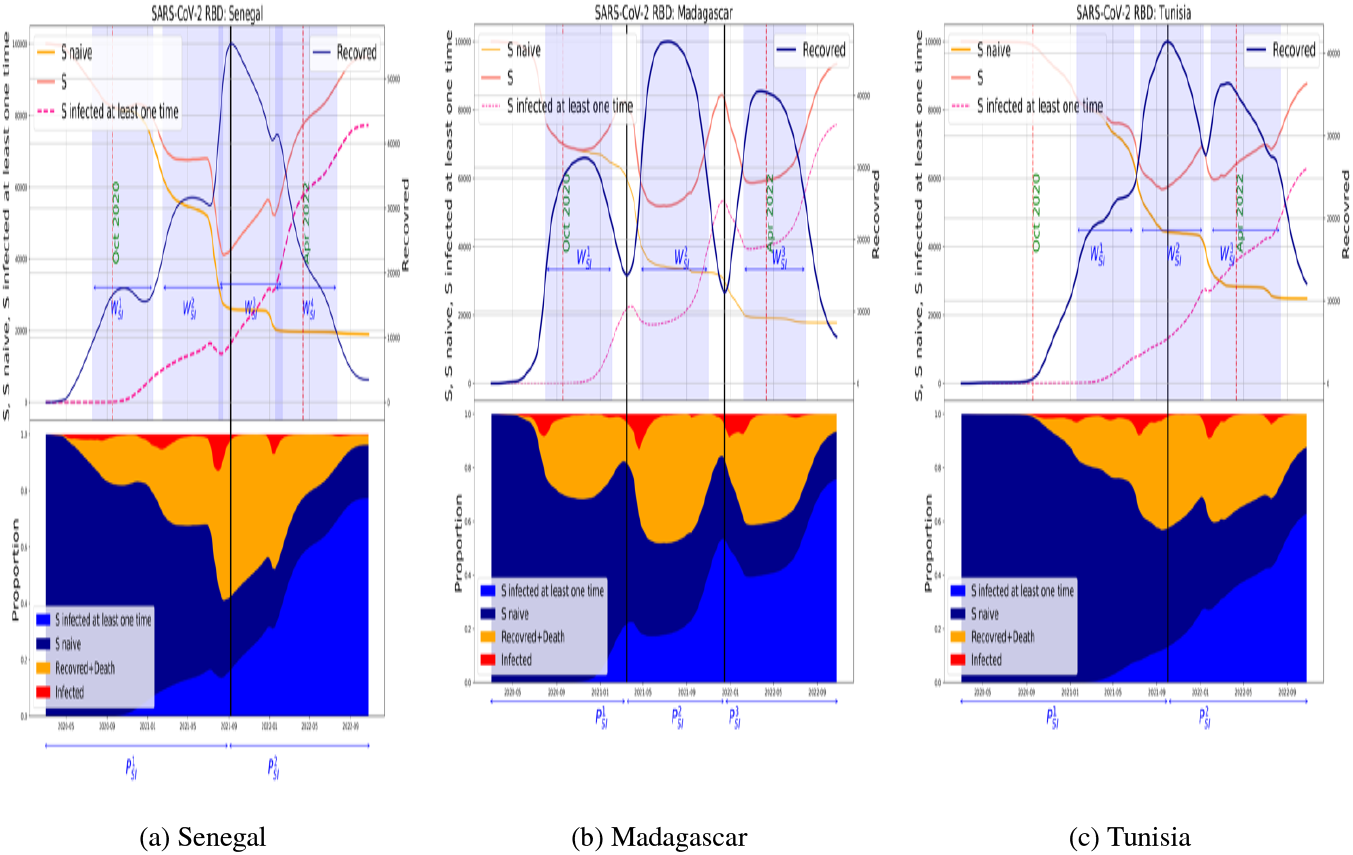
Top: Number of naive susceptibles, susceptibles infected at least once, total susceptibles, and recovered individuals per 100, 000 individuals. Bottom: Same figure as above but in proportion. From left to right: Senegal, Madagascar, and Tunisia. For the three countries, the susceptible naive curve decreases and approaches zero after the third wave. Hence, the infected individuals during waves 1 and 2 are essentially naive. Meanwhile, those infected during waves 3 and 4 are naive and reinfected individuals, respectively. The infected population in wave 4 mostly comprised reinfected individuals. We observed a fluctuation in the curve of susceptible individuals for all waves in Madagascar.

This decrease was more rapid in Senegal than it was in Tunisia, with some observed oscillations in the latter. During the second phase, a radical change was observed in the distribution of individuals within susceptible population, *S*. The number of susceptible/naive individuals, *S*_2_, steeply declined until reaching 25% in Tunisia and 19% in Senegal, approaching zero, whereas the proportion of susceptible/previously infected individuals, S_1_ increased to 63% in Tunisia and 77% in Senegal (Figures 4a, 4c and 5).

In Madagascar, the main feature observed was that the time interval between successive epidemic waves (estimated between 257 and 330 days) was consistently longer than the time duration of the seroimmune waves (estimated between 160, and 173 days). Hence, a new epidemic wave begins when the majority of the individuals who recovered from the preceding epidemic wave have lost their antibodies against SARS-CoV-2. This allowed almost complete reconstitution of the susceptible pool before each new wave (Figures 3b and 3e). In Senegal and Tunisia, the pool of susceptible individuals gradually comprised predominantly those who had been infected at least once, *i*.*e*. primarily composed of individuals in S_1_ category (Figures 4b and 5).

In Senegal, we also computed the protection probability (*i*.*e*. 1–probability of reinfection) for epidemic waves 2–5, 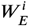 with i in {2, 3, 4, 5}, which corresponded to the ancestral virus, VOC *α* VOC *δ*, and Omicron (Appendix Figure 11). We observed that this probability varied over time during the epidemic wave, with the average of 0.24(σ = 0.09) for the ancestral virus, 0.38(σ = 0.15) for VOC *α*, 0.36(σ = 0.15) for VOC *δ*, and 0.37(σ = 0.17) for the VOC Omicron. The probability of reinfection varied based on the number of individuals who have been previously infected with the virus (*S*_1_) and the duration of their immune response, which in turn depends on the epidemic waves. Recovered individuals may still come into contact with the virus, especially when the epidemic and seroimmune waves intersect or almost intersect, as observed in Senegal and Tunisia. Hence, they can potentially become reinfected, which increases the time spent in the recovered class (*R*) and the probability of reinfection.

Finally, the impact of vaccination on the epidemic curve was assessed by comparing the simulations with and without vaccination in Senegal and Tunisia. Madagascar was not included in the study because vaccine coverage in this country is low (5.75% of the population) (Mathieu et al., 2021). In Tunisia, a massive vaccination campaign began on July 22, 2021, and by February 22, 2022, >12 million doses were administered (58% of the population received at least one dose). This important effort boosted the population’s immunity to SARS-CoV-2, as featured in model simulations.

## 4. Discussion

Modeling the SARS-CoV-2 epidemic in Africa has been difficult owing to limited testing, incomplete data, and varying PHSMs across the continent. To the best of our knowledge, this is the first data-driven SIER model that incorporates immunological data and has been developed using African data. This model simulates the epidemiology of the virus while considering the seroimmune status of the pool of recovered individuals (R) by distinguishing between those who have recovered and maintained a “protective” level of immunity and those who after recovery, have lost this “protection due to the natural antibody decay, thereby shifting back to the pool of susceptibles. This approach had generated curves that accurately depict the onset and amplitude of consecutive epidemic waves, providing valuable insights into the spread of the virus in different African countries.

Previous models have primarily focused on forecasting the impact of interventions (COVID-19 Multi-Model Comparison Collaboration, 2020a,b; Iranzo and Pérez-González, 2021), but only few have incorporated immunity loss; those that have assumed a uniform loss of immunity across the population (Weitz et al., 2020) were unable to evaluate the impact of immune boosting caused by reinfection or vaccination.

The simulation of COVID-19 dynamics in a population structured by antibodies to the S and N virus antigens revealed that anti-N antibodies are mainly indicators of contact with the virus, whereas antibodies to S/RBD can neutralize the virus. Individuals positive for these antibodies seem less susceptible to infection or severe disease. The waning of antibodies over time makes individuals more susceptible to new infections, particularly when new variants emerge. The natural decay of antibodies to S or N antigens after infection was used as a proxy for the decay of immunity against the pandemic virus and its variants. However, this assumption may have limitations, as immunity to SARS-CoV-2 involves diverse mechanisms beyond humoral antibodies response.

In our model, the resumption of a new epidemic wave may have been caused by various factors, including i) re-emergence of the same virus after the reconstitution of a sufficient pool of individuals susceptible to this virus, ii) loss of immunity or relaxation of PHSMs with an increase in human-to-human contact, or iii) epidemic spread in previously non-exposed populations. Alternatively, a new epidemic wave could also be triggered by a new viral variant that has a competitive advantage over other strains, resulting from mutations that increase its fitness or allow it to escape the immunological response.

The fate of COVID-19 as a pandemic will ultimately be determined by the extent to which the immunity induced by the primary infection confers significant protection against reinfection. Stein et al. (2023) have assessed this point through a large meta-analysis and observed high levels of protection (> 82%) against reinfection from ancestral, *α* and *δ* VOCs, However, significantly lower protection (45%) was conferred against reinfection from the omicron BA.1 VOC because of the high immune escape features of this VOC. In the early stages of an epidemic, when a virus emerges within a naive population, the host’s immune responses are not yet constraining, and no selective pressure can yet foster the virus to evolve. Thus, the new virus variants tend to exhibit an increase in the basic reproduction number (*R*_0_). This was evident during the early stages of the pandemic, when the *α* or *β* VOCs of SARS-CoV-2 infected mostly naive populations. These VOCs replicate rapidly in the host and cause serious pathology, with the climax reaching the delta VOC.

However, the increase in *R*_0_ alone cannot account for the intensity of a new wave. The intensity is fueled by the size of the pool of susceptible individuals (*S*), which is continuously replenished by individuals who have lost their immunity after a primary infection and become susceptible to reinfection (*S*_1_). Accordingly, in the three countries examined in the study, the new epidemic waves amplified the emerging VOCs precisely before the seroimmune wave induced by the preceding epidemic wave reached a nadir due to natural antibody decay (in Senegal and Tunisia) or after a longer time interval (in Madagascar).

Starting with the emergence of *δ* VOC, we noted a significant change in the structure of the pool of susceptible. In fact, the ratio of the susceptible still naive to the virus (*S*_2_) to the whole pool of susceptible (*S*) decreased to only 13.5% in Tunisia and 18% in Senegal. At this stage, the virus needs to invest in escaping the immune responses mounted by past infections to support its spread. It also needed to balance its high rate of replication to keep the host sufficiently healthy to infect new ones. This is how the Omicron VOC emerged. In these three countries, the Omicron VOC triggered additional waves with a periodicity varying between 180 and 200 days, *i*.*e*. the time lapse for immunity loss (Dan et al., 2021; Gallais et al., 2021; Gaebler et al., 2021; Ibarrondo et al., 2020).

Factors responsible for the clinical severity of COVID-19 remain elusive and most probably result from a complex interplay of innate and adaptive immune responses as well as population comorbidities (Jordan, 2021). Cell immunity to SARS-CoV-2 acquired after primary infection and reinfection plays an important role as it is more cross-reactive between the different VOCs and attenuates the clinical severity of COVID-19. It would be interesting to extend this work to a model that considers the cellular components of immune responses in order to develop a general model that also predicts the number of severe cases.

Our study demonstrated that the first modest epidemic waves in Tunisia and Senegal possibly resulted from the implementation of stringent PHSMs by health authorities. This significant efforts included widespread testing, contact tracing, and isolation, as well as unprecedented restrictive social measures including closure of businesses, worship and meeting places, ban on public events, intense public information campaigns, and 3-week confinement. In our model, the impact of such measures was monitored in each country using the Oxford SARS-CoV-2 Government Response Stringency Index and the Google Mobility Index.

In developing countries, the implementation of PHSMs had a heavy socio-economic impact, particularly on the impoverished. Therefore, the decreasing compliance of the population with PHSM has led to the re-emergence of the COVID-19 epidemic. Tunisia experienced a deadlier epidemic with 2, 365 deaths per 100, 000 deaths than those in Senegal and Madagascar reporting 113 and 47 deaths, respectively. The factors that may account for these disparities include demographics and individual resistance to severe forms of the disease. However, further studies are required to fully elucidate these factors.

Several surveys in Africa estimated the seroprevalence of SARS-CoV-2 resulting from the successive epidemic waves (Chouikha et al., 2022; Razafimahatratra et al., 2021; Schoenhals et al., 2021; Talla et al., 2022; Uyoga et al., 2021). In Senegal, a national survey performed in November 2021 had revealed that 28.4% of the population have SARSCoV-2 antibodies (Diarra et al., 2022; Talla et al., 2022) while in Tunisia, at the start of the third epidemic wave (March–April 2021) the seroprevalence to SARS-CoV-2 in the general population was 38%. In Antananarivo, Madagascar, SARS-CoV-2 antibody seroprevalence among blood donors was approximately 50% in June 2021 and 70% in October 2021 (Razafimahatratra et al., 2021; Schoenhals et al., 2021). These figures indicated the intensity of the first epidemic wave and rapid decay of SARS-CoV-2 antibodies in Madagascar. During these two periods, the percentage recovery increased from approximately 40% in June to 50% in October (Figure 5).

**Figure 5:**
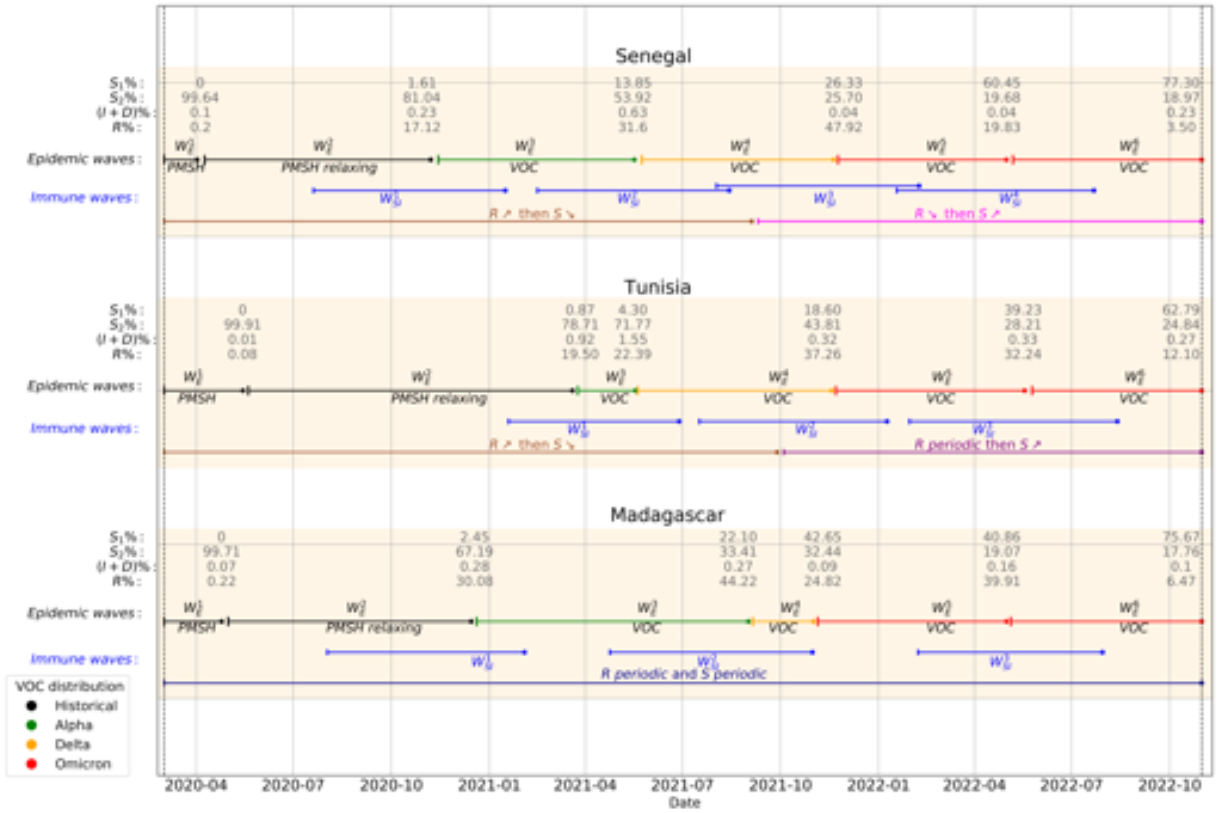
Summary of different epidemiological and seroimmune waves with their intensities and VOCs. The percentiles of the pool of susceptible, *S*_1_ and S_2_, recovered, *R*, and infected and died, *I* + *D*, individuals at the beginning of each wave are also presented. The annotation below of the epidemiological waves represents the reason for the wave: PHSM, wave regulated by PHSM; PHSM relaxing, wave caused by PHSM relaxing; and VOC, wave caused by the introduction of a new VOC. Senegal, Tunisia, and Madagascar experienced six epidemiological waves caused by the same event and VOC. However, the appearance of these waves creates different seroimmune responses. They intersect in Senegal, are very close to each other in Tunisia, and are very distinct in Madagascar. This spacing makes the number of recoveries and susceptibility periodic in Madagascar, which implies that the waves can be considered independent. This was not the case in Senegal or Tunisia.

Finally, we demonstrated that the introduction of anti-SARS-CoV-2 vaccination had a significant effect in Tunisia, but a very modest effect in Senegal, reflecting the respective intensity of the vaccination effort in each country.

## 5. Conclusion

This study provides valuable insights into the dynamics of the SARSCoV-2 pandemic in African countries, with a focus on the impact of immunity loss. Utilizing a SEIR/DS mathematical model, we monitored the seroimmune status of individuals over a 3-year period in Tunisia, Senegal, and Madagascar, representing diverse African contexts. These findings shed light on the evolving nature of the epidemic and its association with immune dynamics.

In the present study, we observed in Senegal and Tunisia, the epidemic followed a pattern characterized by three distinct periods, with recurring waves attributed to immunity loss against emerging VOCs. In contrast, in Madagascar, the interval between successive waves was consistently longer than the time required for immunity loss. By the end of the *δ* and Omicron epidemic waves, a substantial proportion of the population in all three countries had been infected by SARS-CoV-2. These findings are particularly significant when considered alongside recent research demonstrating that previous infections provide a high level of protection against severe diseases caused by VOCs *α, β* and *δ*, as well as Omicron BA.1, for up to 40 weeks. This optimistic outlook, coupled with the widespread infection rates observed in the study countries, suggests a potentially favourable trajectory for future waves of the pandemic in Africa.

## Data Availability

All data produced in the present study are available upon reasonable request to the authors

## Ethical Approval statement

This research project involves the use of anonymized population-level data.

## CRediT authorship contribution statement

**Bechir Naffeti:** Formal analysis/Model construction, Simulation and visualisation. **Walid BenAribi:** Formal analysis/Model construction. **Amira Kebir:** Formal analysis/Model construction, Review and editing. **Maryam Diara:** Review and editing. **Matthieu Schoenhals:** Conceptualisation, Review and editing, Data provider. **Inès Vigan-Womas** : Conceptualisation, Review and editing, Data provider. **Koussay Dellagi:** Conceptualisation, Research conducting, Review and editing. **Slimane BenMiled:** Conceptualisation, Formal analysis/Model construction, Simulation and visualisation, Research conducting, Writing the first draft, Review and editing, Data provider.

## Footnotes

1 https://covid19.who.int

2 https://www.indexmundi.com

3 https://covid19.who.int

4 https://www.google.com/covid19/mobility,

5 https://ourworldindata.org/covid-google-mobility-trends

