## supplementary for "Comparative Reconstruction of SARS-CoV-2 transmission in three African countries using a mathematical model integrating immunity data"

### 1. Appendix: Model assumptions

The model assumptions are as follows:

1. We consider that the population is subdivided into Susceptible, Exposed, Infected and Recovered. The Susceptible are further subdivided into two groups: $S_1$ (individuals that were previously infected but have lost their antibodies to S/RBD) and susceptible $S_2$ who are SARS-CoV-2 naive.
2. Only Exposed or Infected individuals can transmit the disease. Recovered cannot transmit the disease as long as they have antibodies to S/RBD (*i.e.,* remain immune). The force of the infection is proportionate to the weighted sum of the infected ($I_d$ and $I_{nd}$ ) and the and exposed, $E$, population density (frequency-dependence hypothesis) [5, 4].
3. An Individual in the Recovered class shifts to the pool $S_1$ of Susceptible ones once he loses antibodies to S/RBD.
4. Vaccination takes Susceptible or Recovered individual into the Recovered class with maximum sero-immune level.
5. An individual in the Recovered class can be re-infected and if so the re-infection boosts its sero-immunity to a higher level.
6. The sero-immune loss function depends only on the individual sero-immune level (no inter-variability between individual with the same level of the sero-immune state) and is either a linear regression, or a logistic regression.

### 2. Appendix: Model description

We describe the full dynamics of SARS-CoV-2 infection by extending the model developed by [7, 2] to SEIR/DS (Susceptible $S$, Exposed $E$, Infected $I$, Recovered $R$, and Death due to disease $D$), see figure 1. The compartment of Susceptible, $S$, is subdivided into two groups: those denoted $S_1$, who were previously infected at least one time by SARS-CoV-2, and those denoted $S_2$ who are naive to the virus. The susceptible $S$ individuals are infected when they interact with infected pre-symptomatic $E$ (yet infectious) or infected symptomatic individuals, $I$ (hypothesis 2). As described by the mass action law, the susceptible individuals become infected and move to the class $E$ at a rate proportional to their exposure to infected individuals, $S_i(t)\beta\left(\frac{E(t)+I_{nd}(t)+I_d(t)}{N(t)}\right)$, $i = 1, 2$, with $\beta$ the contact rate. An exposed individual develops symptoms after an incubation period, $\frac{1}{\delta}$ days, and then moves to the Infected class. The infected compartment is divided into two groups: those who have not been detected $I_{nd}$ and those who have been detected $I_d$. In this work, we assume that the number of reported cases at each time $t$ is proportional to the number of all cases for each time $t$. Let's denote the proportion coefficient by $\alpha$. Therefore, the rate of asymptomatic infectious becoming reported symptomatic is $\alpha\delta$ and the rate of the unreported cases is $(1-\alpha)\delta$. Finally, an infected individual is dead after $\frac{1}{\sigma}$ days with probability $\epsilon$, or recovered with probability $(1-\epsilon)$ and goes to class $R$. OOver a period of time, the individual goes through different sero-immune levels - and then becomes susceptible to infection again.

Let $r(t,\tau)$ be the density of immune individuals at time $t$ with an sero-immune level $\tau \in [\tau_{\min}, \tau_{\max}]$, where $\tau_{\min}$ corresponds to a low sero-immune level and $\tau_{\max}$ corresponds to the maximal sero-immune level. Thus, the total number of recovered individuals at time $t$ is given by::

$$R(t) = \int_{\tau_{min}}^{\tau_{max}} r(t,\tau)d\tau.$$

Individuals who recover at time $t$ enter the immune compartment $R$ with a maximum sero-immune level $\tau_{\max}$. Their sero-immune level then decreases at the rate $f(\tau)$ until it reaches the minimum value $\tau_{\min}$, where they become susceptible again. We suppose that the sero-immune loss rate $f(\tau)$ is constant for all individuals with the same sero-immune level $\tau$ (hypothesis 6). Therefore, the sero-immune loss equation is:

$$\frac{d\tau(t)}{dt} = -f(\tau) \text{ with } \tau \in [\tau_{min}, \tau_{max}].$$

In the absence of immune system boosting, an infected individual who recovered at time $t$ becomes again susceptible at time $t+T$, where:

$$T = \int_{\tau_{\min}}^{\tau_{max}} \frac{1}{f(\tau)} d\tau.$$

Let $d\tau$ be a short increase of sero-immune level and $[\tau - d\tau, \tau]$ a small sero-immune interval. We have, at time $t$, over a short time step $dt << 1$, the number of individuals entering and leaving $[\tau - d\tau, \tau]$ is $r(t, z)dt$.

As sero-immune level tends to decrease over time at the rate $f(\tau)$ (hypothesis 6), individuals enter the interval from $\tau$ and exit from $\tau - d\tau$. Thus, per unit of time, $dt$, the inflows in the interval $[\tau - d\tau, \tau]$ are equal to $r(t,\tau)f(\tau)dt$, and the outflows are equal to $r(t, \tau - d\tau)f(\tau - d\tau)dt$.

However, the sero-immune system can be strengthened by vaccination or contact with an infected person, allowing individuals with sero-immune level $[\tau - d\tau, \tau]$ to switch to a higher level, and those with lower sero-immune levels to "jump" to the interval $[\tau - d\tau, \tau]$ (hypothesis 4 and 5).

We assume that vaccination jumps the sero-immune level to $\tau_{max}$ (hypothesis 4). Let $\upsilon_3$ be the vaccination rate of recovery, and the probability that a vaccinated individual at level $\tau$ increases his sero-immune level to $\tau_{max}$ is $p_\upsilon(\tau)$. The outflow from the sero-immune interval $[\tau - d\tau, \tau]$ due to vaccination is $\upsilon_3 p_\upsilon(\tau)r(t,\tau)d\tau dt$. This outflow is part of the influx of $r(t, \tau_{max})$ (boundary condition).

We assume that the recovered individuals cannot transmit the disease as long as they have antibodies to S/RBD (hypothesis 2). The force of infection is proportional to the weighted sum of the infected ($I_n$ and $I_{nd}$) and the exposed, $E$, population density (frequency-dependence hypothesis). Therefore, the force of infection is $\beta\frac{E+I_d+I_{nd}}{N}$.

For all $\upsilon \in [\tau_{min}, \tau_{max}]$ such that $\upsilon \leq \tau$, let $p(\tau, \upsilon)$ denote the probability that an individual with sero-immune level $\upsilon$ moves to sero-immune level $\tau$ when exposed to the pathogen (hypothesis 5). Let's extend this definition for all $\upsilon \in ]\tau, \tau_{max}]$ by setting $p(\tau, \upsilon) = 0$. Thus, per unit of time, $dt$, the entries in the interval $[\tau - d\tau, \tau]$ by infection are equal to,

$$\beta\frac{E + I_d + I_{nd}}{N}\left(\int_{\tau_{min}}^{\tau_{max}} r(t,\upsilon)\ p(\tau,\upsilon)d\upsilon\right)\ d\tau\ dt,$$

and the outgoing individual are equal to, as $\int_{\tau_{min}}^{\tau_{max}} p(\upsilon,\tau)d\upsilon = 1$:

$$\beta\frac{E + I_d + I_{nd}}{N} r(t,\tau)d\tau dt.$$

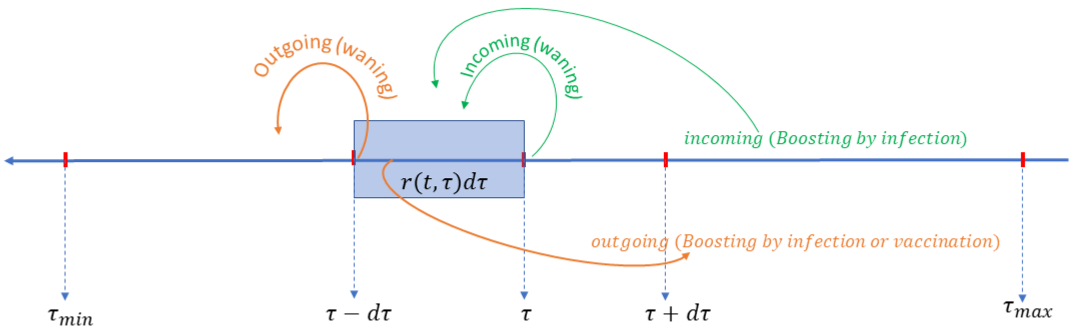


**Figure 1:** Conceptual model

Therefore, the total number of immune individuals with sero-immune level in $[\tau - d\tau, \tau]$ at time $t + dt$ is:

$$\begin{aligned} r(t+dt,\tau)d\tau = &\underbrace{r(t,\tau)d\tau}_{\text{at time } t} - \underbrace{r(t,\tau-d\tau)f(\tau-d\tau)dt}_{\text{outgoing (waning)}} + \underbrace{r(t,\tau)f(\tau)dt}_{\text{incoming (waning)}} - \underbrace{\beta\frac{E+I_d+I_{nd}}{N}r(t,\tau)d\tau dt}_{\text{outgoing (Boosting by infection)}} \\ &\underbrace{+\beta\frac{E+I_d+I_{nd}}{N}\sum_{\upsilon\in G_\tau} r(t,\upsilon)\ d\upsilon\ p(\tau,\upsilon)\ d\tau\ dt}_{\text{incoming (Boosting by infection)}} - \underbrace{\upsilon_3 p_\upsilon(\tau)r(t,\tau)d\tau dt}_{\text{outgoing (Boosting by vaccination)}} \end{aligned} \tag{1}$$

Dividing equation (1) by $d\tau$ and then making $d\tau$ tends towards 0, we obtain:

$$\begin{aligned} r(t+dt,\tau) - r(t,\tau) = {} & dt\frac{\partial}{\partial\tau} r(t,\tau) f(\tau) - \beta\frac{E(t)+I_d(t)+I_{nd}(t)}{N} r(t,\tau)dt \\ & + dt\beta\frac{E(t)+I_d(t)+I_{nd}(t)}{N}\int_{\tau_{min}}^{\tau} p(\tau,v)r(t,v)dv - v_3 p_v(\tau) r(t,\tau)dt \end{aligned} \tag{2}$$

Dividing equation (2) by $dt$ and then making $dt$ tends towards 0, we obtain:

$$\begin{aligned} \frac{\partial}{\partial t} r(t,\tau) = {} & \frac{\partial}{\partial\tau} r(t,\tau) f(\tau) - \beta\frac{E(t)+I_d(t)+I_{nd}(t)}{N} r(t,\tau) \\ & + \beta\frac{E(t)+I_d(t)+I_{nd}(t)}{N}\int_{\tau_{min}}^{\tau} p(\tau,v)r(t,v)dv - v_3 p_v(\tau) r(t,\tau) \end{aligned} \tag{3}$$

The boundary condition is constitute of individual who are infected and not died, susceptible vaccinated, recovery that were vaccinate and recovery that get infected with a boost that make their sero-immune level to go to $\tau_{max}$. Therefore we have:

$$\begin{aligned} f(\tau_{max}) r(t,\tau_{max}) = {} & ((1-\epsilon)\sigma(I_d(t)+I_{nd}(t)) + v_1 S_1(t) + v_2 S_2(t)) \\ & + \beta\frac{E(t)+I_d(t)+I_{nd}(t)}{N}\int_{z_{min}}^{z_{max}} r(t,v)\, p(\tau_{max},v)dv \\ & + v_3\int_{z_{min}}^{z_{max}} p_v(v) r(t,v)dv \end{aligned} \tag{4}$$

Based on the above equations, the epidemiological model is as following:

$$\begin{cases} \dot{S}_1(t) &= -\beta\left(\dfrac{E(t)+I_{nd}(t)+I_d(t)}{N(t)}\right) S_1(t) - v_1 S_1(t) + f(\tau_{min}) r(t,\tau_{min}) \\ \dot{S}_2(t) &= -\beta\left(\dfrac{E(t)+I_{nd}(t)+I_d(t)}{N(t)}\right) S_2(t) - v_2 S_2(t) \\ \dot{E}(t) &= \beta\left(\dfrac{E(t)+I_{nd}(t)+I_d(t)}{N(t)}\right)(S_1(t)+S_2(t)) - \delta E(t) \\ \dot{I}_{nd}(t) &= \alpha\delta E(t) - \sigma I_{nd}(t) \\ \dot{I}_d(t) &= (1-\alpha)\delta E(t) - \sigma I_d(t) \\ \dot{D}(t) &= \epsilon\sigma(I_{nd}+I_d) \end{cases} \tag{5}$$

Where $S_1(0) = 0$, $S_2(0) = S_0 > 0$, $E(0) = E_0 \geq 0$, $I_{nd}(0) = I_{nd_0} \geq 0$, $I_d(0) = I_{d_0} \geq 0$, $D(0) = 0$ and it is coupled with the following partial differential equation (PDE):

$$\begin{cases} \dfrac{\partial}{\partial t} r(t,\tau) - \dfrac{\partial}{\partial\tau}(f(\tau) r(t,\tau)) & = \beta\left(\dfrac{E(t)+I_{nd}(t)+I_d(t)}{N(t)}\right)\left(\displaystyle\int_{\tau_{min}}^{\tau} p(\tau,u) r(t,u)du - r(t,\tau)\right). \\ & \quad -v_3 p_v(\tau) r(t,\tau) \\ f(\tau_{max}) r(t,\tau_{max}) & = ((1-\epsilon)\sigma(I_d(t)+I_{nd}(t)) + v_1 S_1(t) + v_2 S_2(t)) \\ & \quad +\beta\dfrac{E(t)+I_d(t)+I_{nd}(t)}{N}\displaystyle\int_{\tau_{min}}^{\tau_{max}} r(t,v)\, p(\tau_{max},v)dv \\ & \quad +v_3\displaystyle\int_{\tau_{min}}^{\tau_{max}} p_v(v) r(t,v)dv \\ r(0,\tau) & = 0 \end{cases} \tag{6}$$

**Table 1**
Epidemiological and demographic data

| Parameter | Description | Country | | | Ref. |
|---|---|---|---|---|---|
| | | Tunisia | Madagascar | Senegal | |
| $N$ | Population size | 11818619 | 26970000 | 16705608 | [1] |
| $1-\alpha$ | Fraction of undeclared infectious | 0.7 | 0.7 | 0.7 | |
| $v_1$, $v_2$ | Vaccination rate of individuals naive | 0.0015 | 0.00012 | 0.0001 | [2] |
| $\delta$ | Incubation rate | 1/5 | 1/5 | 1/5 | [6] |
| $\beta$ | Contact rate | estimate every 10 days | | | |
| $\sigma$ | Recovered rate | 1/10 | 1/10 | 1/10 | [3] |
| $\epsilon$ | Death rate caused by the virus | 0.0025 | 0.00006 | 0.0001 | [3] |

1. www.indexmundi.com/
2. covid19.who.int/
3. ourworldindata.org/mortality-risk-covid/

**Table 2**
Linear Regression for sero-immune loss kinetics.

| | $\tau_{min}$ | $\tau_{max}$ | a | b | R2 | $P > \|t\|$ | T (days) |
|---|---|---|---|---|---|---|---|
| RBD, 180 days | 0.007 | 0.19 | −0.001 | 0.19 | 0.8 | 0.04 | 183 |
| RBD scenario 210 days | 0.007 | 0.19 | −0.0007 | 0.19 | 0.8 | 0.018 | 210 |
| NDVET | 0.43 | 0.56 | −0.0006 | 0.55 | 0.8 | 0.01 | 200 |

### 3. Appendix: Results

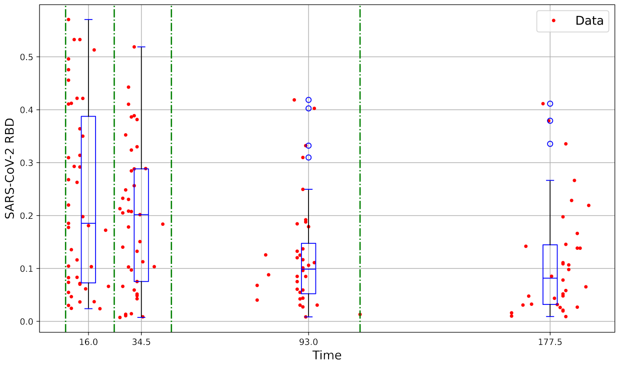


(a) RBD/S data for kinetic

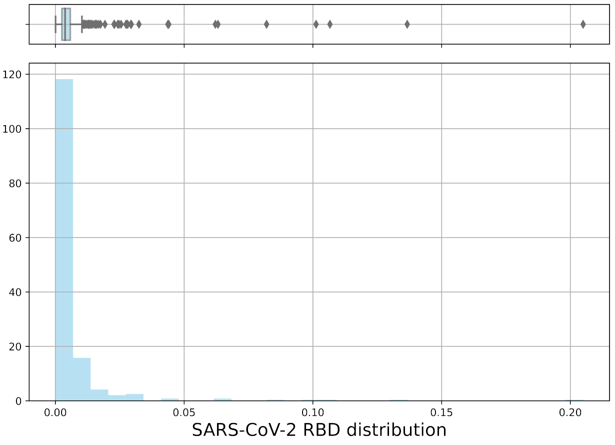


(b) RBD/S prepandemic data

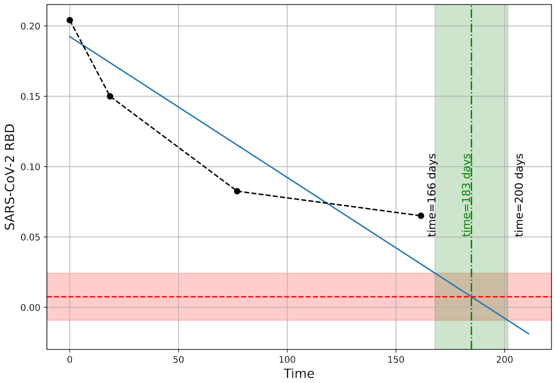


(c) RBD/S linear kinetic model

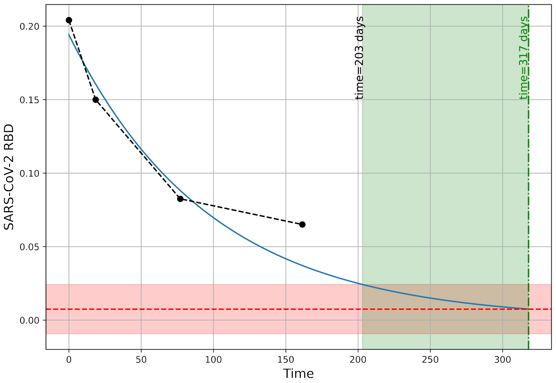


(d) RBD/S non-linear kinetic model

**Figure 2:** Kinetic and prepandemic antibodies to the RBD/S protein Data distribution (sub-figure 2a and 2b). Sub-figure 2c and 2d: Linear and Nonlinear kinetic model, $f(t)$. We used four data sets acquired in Senegal between August 14, 2020 and August 17, 2021 (corresponding to epidemic waves 2, 3, and 4). Data were normalized by technology and by protein. We divided the normalized kinetic data into four-time groups: $>$ 150 days; [50,150] days; [23,50] days and 23 days and performed a linear and a nonlinear regression.

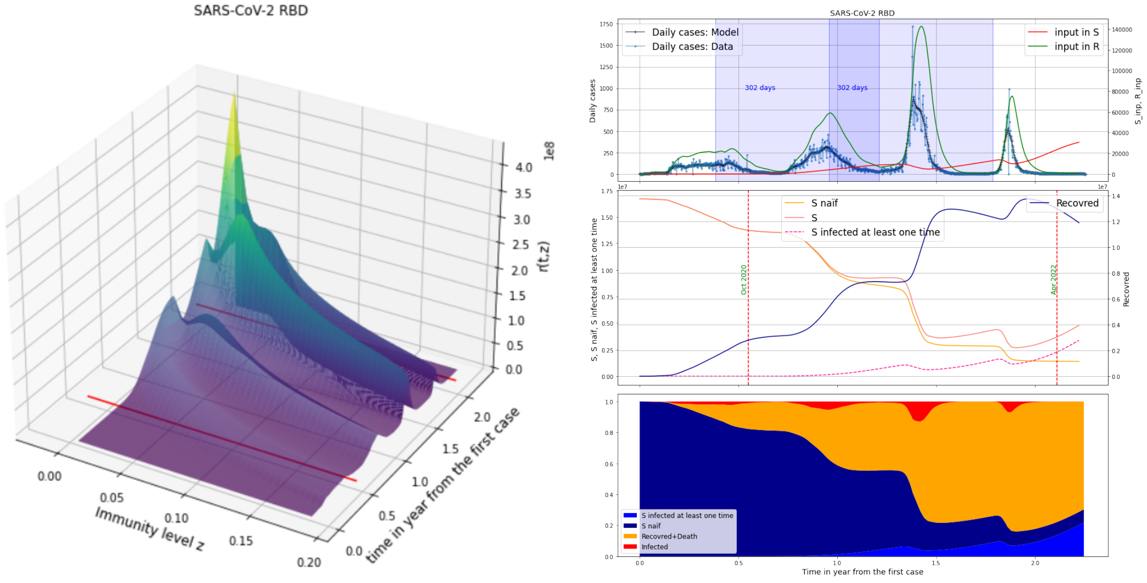


(a) Senegal $r(t, \tau)$

(b) Senegal $r(t, \tau)$ cross-section

**Figure 3:** From left to right: Figures 4a, 4c, 4e represent the simulation data of $r(t, \tau)$ from the begin of the epidemic to September 2022, with $\tau$ the amount of IDVet (IgG anti-N protein). Figures 4b, 4d, 4f represent the cross-section of $r(t, \tau)$ for $\tau = \tau_{max}$ ($R$ input class) and $\tau = \tau_{min}$ ($S$ input class) as well as the curve of the daily case by time (in days) with daily case field data.

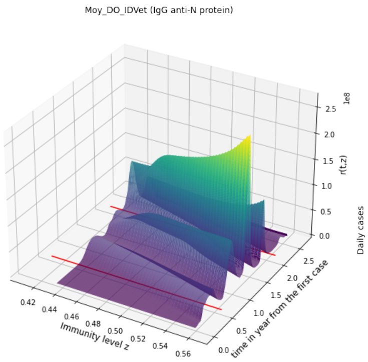


(a) Senegal $r(t, \tau)$

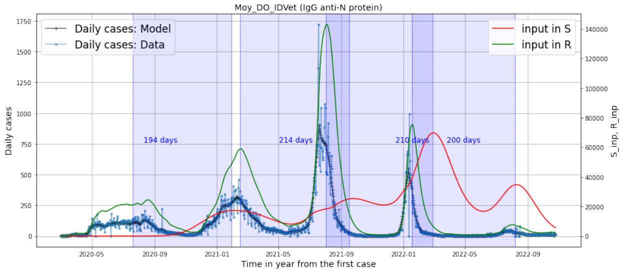


(b) Senegal $r(t, \tau)$ cross-section

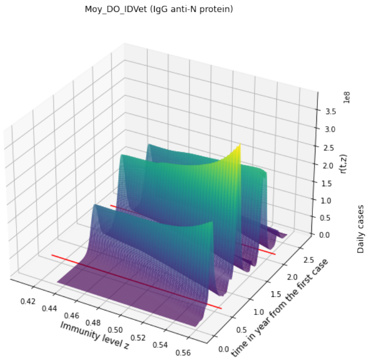


(c) Madagascar $r(t, \tau)$

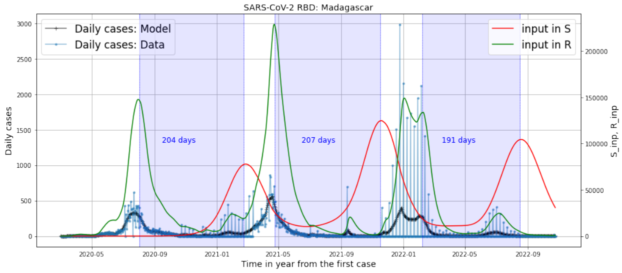


(d) Madagascar $r(t, \tau)$ cross-section

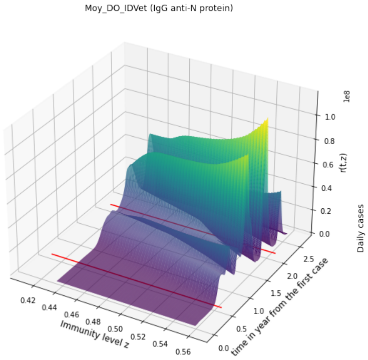


(e) Tunisia $r(t, \tau)$

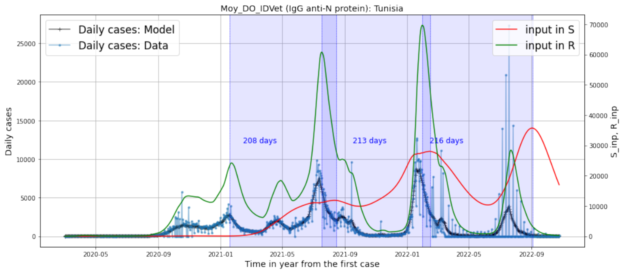


(f) Tunisia $r(t, \tau)$ cross-section

**Figure 4:** From left to right: Figures 4a, 4c, 4e represent the simulation data of $r(t, \tau)$ from the begin of the epidemic to September 2022, with $\tau$ the amount of IDVet (IgG anti-N protein). Figures 4b, 4d, 4f represent the cross-section of $r(t, \tau)$ for $\tau = \tau_{max}$ ($R$ input class) and $\tau = \tau_{min}$ ($S$ input class) as well as the curve of the daily case by time (in days) with daily case field data.

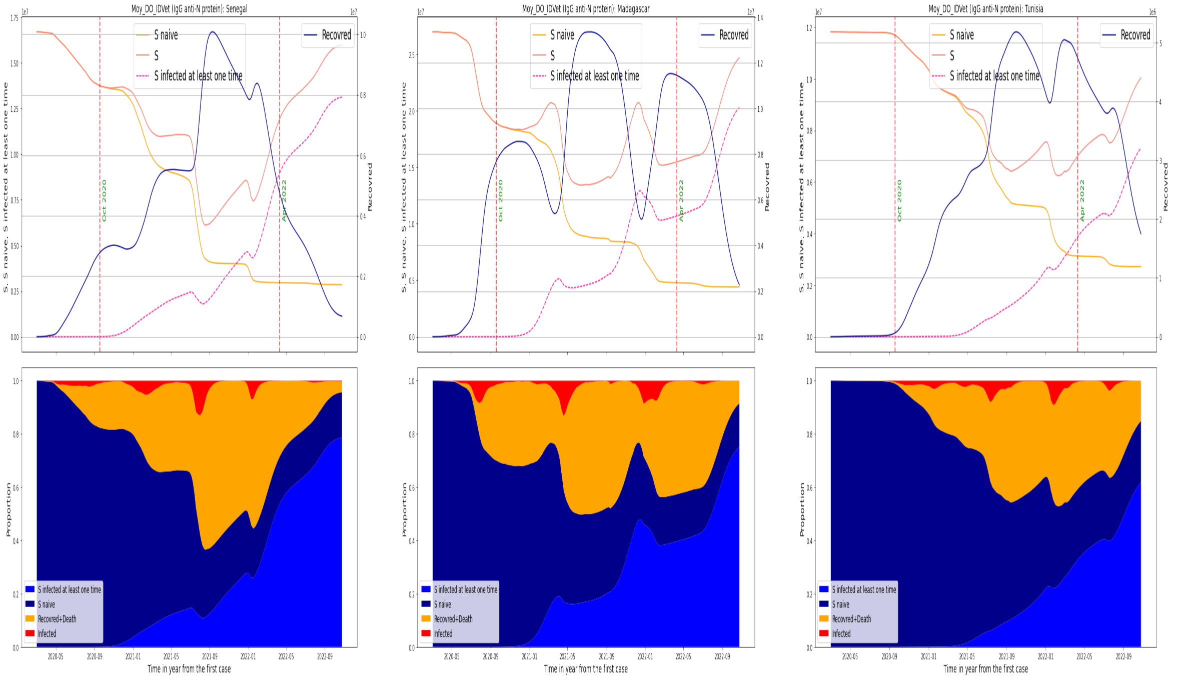


(a) Senegal

(b) Madagascar

(c) Tunisia

**Figure 5:** Bottom: Number of naive susceptible, susceptible infected at least ones, total susceptible and recovered amount of IDVet (IgG anti-N protein). Same figure than above but in proportion. From left to right: Senegal, Madagascar, and Tunisia. For the three countries, we observe that the susceptible naive curve decreases and tends in percentage to 0 after the third wave. Hence, the infected of waves 1 and 2 are essentially naive. On the other hand, those of waves 3 and 4 come from naive people and re-infected people. Wave 4 is mostly made up of re-infected people. We notice a fluctuation in the curve of susceptible individuals for all waves in Senegal and from wave 4 in Tunisia. This implies independence of the waves in relation to each other.

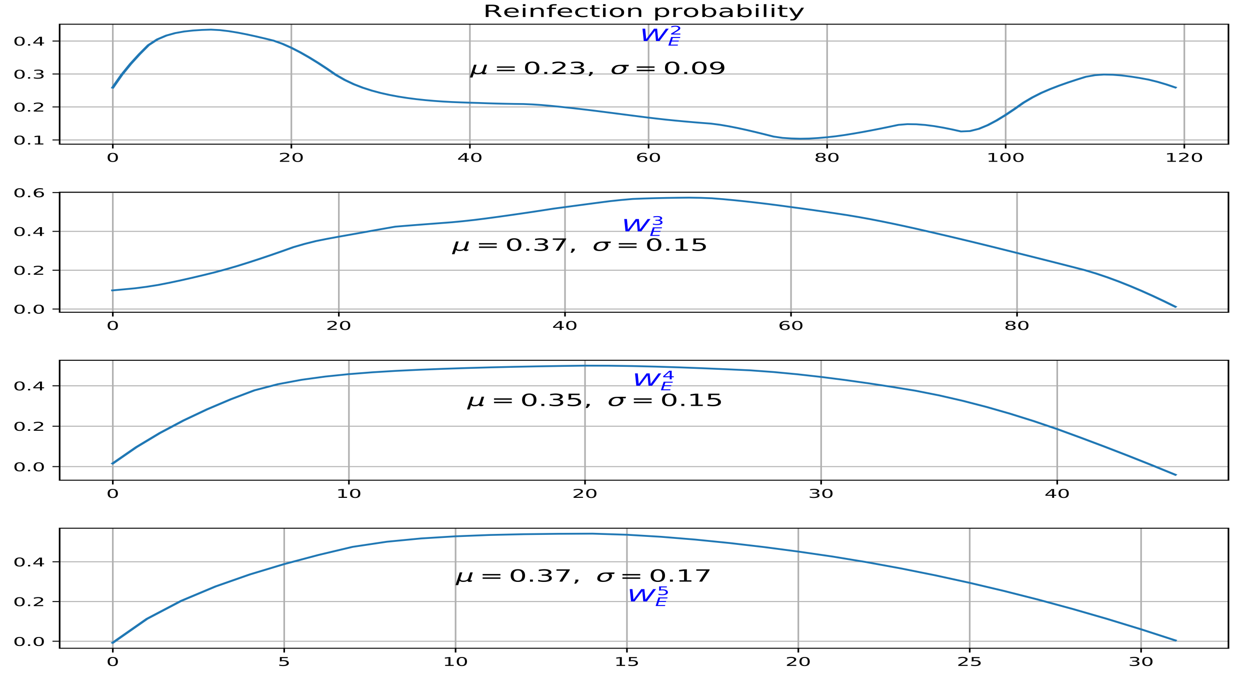


**Figure 6:** Probability of protection against reinfection (*i.e.*, 1− probability of reinfection) in the case of Senegal and for $W_E^i$ with $i \in \{2, 3, 4, 5\}$ epidemic waves $2-5$, caused by the historical strain, $\alpha$, $\delta$ and Omicron VOCs

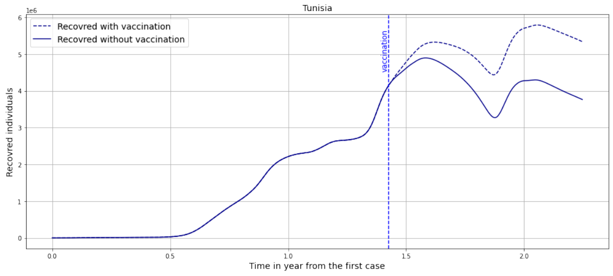


(a) Tunisia

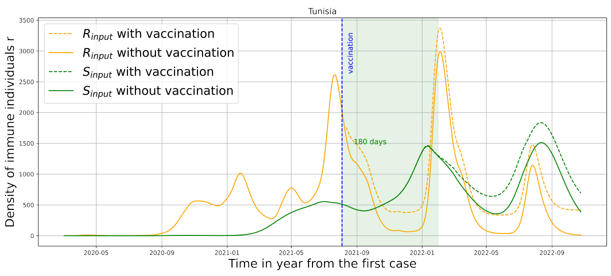


(b) Tunisia

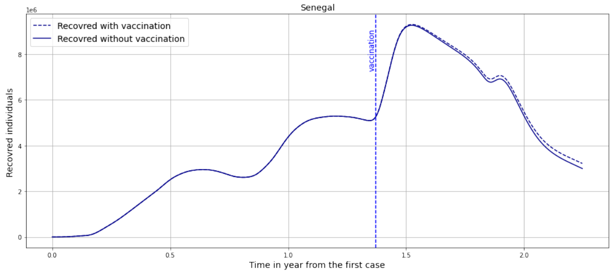


(c) Senegal

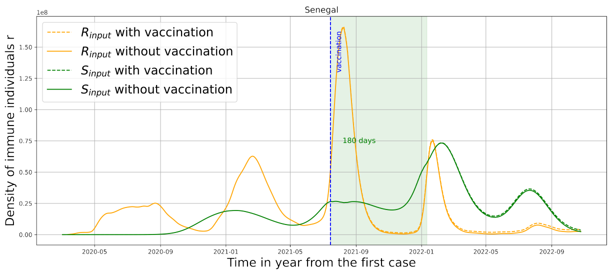


(d) Senegal

**Figure 7:** Simulation of the effect of vaccination on the Recovery class (Fig 7a) and on the input of the recovery class and the input of the susceptible class (fig 7b) in Tunisia.

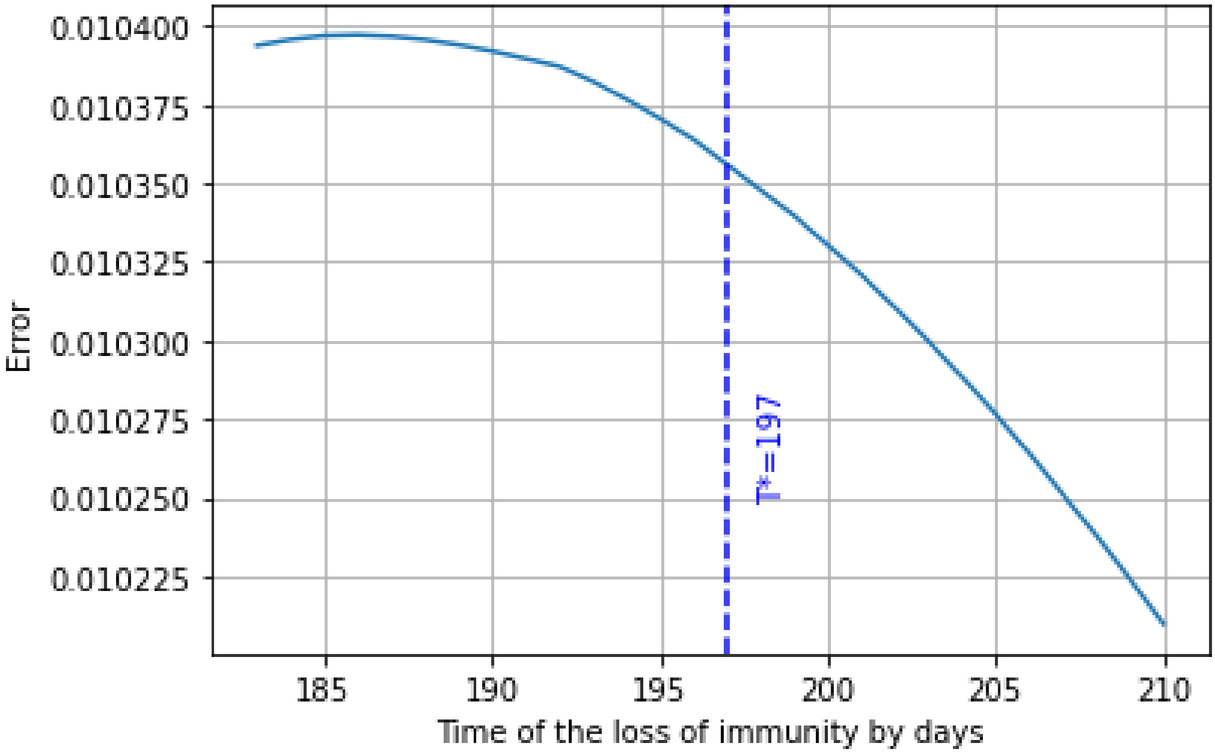


**Figure 8:** Sensitivity analysis of $r(t,z)$ on time required for immunity loss, $T$, between $180$ and $210$ days.
